# Perceptions of COVID-19 risk, vaccine access, and confidence: a qualitative analysis of South Asians in Canada

**DOI:** 10.1101/2022.10.21.22281321

**Authors:** Sujane Kandasamy, Baanu Manoharan, Zainab Khan, Rosain Stennett, Dipika Desai, Rochelle Nocos, Gita Wahi, Davina Banner, Russell J. de Souza, Scott A. Lear, Sonia S. Anand

**Affiliations:** Department of Health Research Methods, Evidence & Impact, McMaster University, Hamilton, Ontario, Canada; Master of Public Health Program, McMaster University, Hamilton, Ontario, Canada; Population Health Research Institute, McMaster University, Hamilton, Ontario, Canada; Community Health Research Team (CoHeaRT), Simon Fraser University at St. Paul’s Hospital, Burnaby, British Columbia, Canada; Department of Pediatrics, McMaster University, Hamilton, Ontario, Canda; School of Nursing, University of Northern British Columbia, Prince George, British Columbia, Canada; Faculty of Health Sciences, Simon Fraser University, Burnaby, British Columbia, Canada; Department of Medicine, McMaster University, Hamilton, Ontario, Canada

**Keywords:** COVID-19, vaccine uptake, South Asian health, qualitative methodologies, interviews

## Abstract

**VISUAL ABSTRACT CAN BE FOUND <u>HERE</u>**

**Objectives:** In the first full year of the COVID-19 pandemic (2020), South Asians living in the Greater Toronto Hamilton and Vancouver Areas experienced specific barriers to accessing SARS-CoV-2 testing and receiving reliable health information. However, between June 2021 and February 2022, the proportion of people having received at least 1 dose of a COVID-19 vaccine was higher among this group (96%) than among individuals who were not visible minorities (93%). A better understanding of successful approaches and the challenges experienced by those who remain unvaccinated among this highly vaccinated group may improve public health outreach in subsequent waves of the current pandemic or for future pandemic planning. Using qualitative methods, we sought to explore the perceptions of COVID-19 risk, vaccine access, uptake, and confidence among South Asians living in Canada.

**Methods:** In this qualitative study, we interviewed 25 participants between July 2021 and January 2022 in the Greater Toronto Hamilton and Greater Vancouver Areas (10 community members, 9 advocacy group leaders, 6 public health staff). We conducted initial and focused coding in duplicate and developed salient themes. Throughout this process, we held frequent discussions with members of the study’s advisory group to guide data collection as it relates to community engagement, recruitment, and data analysis.

**Results:** Access to and confidence in the COVID-19 vaccine was impacted by individual risk perceptions; sources of trusted information (ethnic and non-ethnic); impact of COVID-19 and the pandemic on individuals, families, and society; and experiences with COVID-19 mandates and policies (including temporal and generational differences). Approaches that include community-level awareness and tailored outreach as it relates to language and cultural context were considered successful.

**Conclusion:** Understanding factors and developing strategies that build vaccine confidence can guide our approach to increase vaccine acceptance in the current and future pandemics.

## Introduction

The SARS-COV-2 (COVID-19) global pandemic, officially declared in March 2020, abruptly and profoundly altered home and work life for Canadians world-wide (Cucinotta & Vanelli, 2020). The COVID-19 pandemic has resulted in unprecedented challenges across society in Canada, including the surge of “parallel pandemics” related to inequities (Ismail et al., 2021), mental health (Yao et al., 2020), and chronic disease (Kendzerska et al., 2021). During the period of the pandemic where vaccination was not available, access to testing and adaptation to different ways of working predominated risk reduction efforts. COVID-19 vaccine development began in January 2020 and advanced rapidly. However, factors related to equity and individual interest/acceptability of vaccines influences. “Vaccine confidence” is a term that encompasses a spectrum of beliefs and attitudes towards vaccines. Low vaccine confidence is characterized by uncertainty and ambivalence about vaccination impact and uptake (Liao et al., 2019). Surveys conducted in high-income countries have shown there is greater hesitancy regarding the COVID-19 vaccine among people from ethnic minorities (Robinson et al., 2021). These findings parallel a historical trend of lower vaccine uptake in areas with higher proportions of ethnic minority communities (UK Government Scientific Advisory Group for Emergencies, 2020).

The South Asian community in Canada refers to people who originate from the Indian sub-continent. South Asians represent the second largest, but fastest growing non-white ethnic group in Canada (Census Profile, 2016). South Asian communities in Ontario (ON) and British Columbia (BC) were and are especially impacted by the pandemic. The higher infection rates from COVID-19 within South Asian communities is largely explained by socio-cultural factors, including occupational factors (e.g., essential work) moreso than biological differences (e.g., genetic predisposition, pathophysiological differences in susceptibility or response to infection) (Anand SS et al CMAJ Open McKenzie & Petersen, 2021).

Public health measures, including testing, mask mandates, quarantine measures, restrictions, and vaccination, were implemented and promoted by federal, provincial, and municipal governments, as well as local public health units. Most messaging regarding these measures were not tailored for different cultural communities, even though Canada is home to numerous non-White cultural communities. This may represent a missed opportunity to engage with these communities, but more importantly, a failure to get buy-in/uptake, eroding public trust.

We established the COVID CommUNITY study, a prospective cohort study focused on South Asian adults living in Canada (Ontario and British Columbia) to better explore COVID-19 vaccine immune response, safety, and confidence (Anand et al., 2022). We wanted to deepen our understanding of the factors that influenced COVID-19 vaccine confidence by braiding together South Asian community members, advocacy group leaders, and public health staff perspectives of vaccine access and uptake.

## Design and Methods

This qualitative study aimed to explore the perceptions of risk regarding COVID-19 infection and its consequences, and the perceptions of vaccine access and uptake among South Asian individuals, advocacy group leaders, and public health staff in Canada. All participants provided informed consent and this study was approved by Hamilton Integrated Research Ethics Board (13323 - March 24, 2021) and the British Columbia Research Ethics Board (H21-00866 – June 18, 2021).

A Qualitative Descriptive approach (QD) was adopted as a method of naturalistic inquiry that uses low inference interpretation (Clarke & Braun 2013; Sandelowski., 2010). The goal of QD is to develop and report an accurate account of participant experiences, events, and process (Braun & Clarke, 2006). Using a thematic analysis approach, the five members of the analysis team (SK, BM, ZK, DD, RS) read the transcripts multiple times to become familiar with the data, generate initial codes, combine codes into overarching themes, hold discussions to clarify ideas, and develop a robust description of the findings (Clarke et al., 2014).

### Setting

Our study focused on South Asian community members, advocacy group leaders, and public health staff who live or work in the Greater Toronto Hamilton Area (GTHA) and Greater Vancouver Area (GVA). We enrolled participants from the two largest urban South Asian communities in Canada, with the makeup of these communities being representative of the broader Canadian immigration landscape. We recruited broadly within these geographical settings with the support of community partners (e.g., vaccine centers, mobile vaccine clinics, university campuses, local organizations).

### Process

Once a participant expressed interest, they were informed of the study procedures. If they wished to participate, informed consent was obtained, and a virtual interview was scheduled (either via Zoom or telephone depending on the participant’s preference). Interviews, conducted by either SK (ON) or RN (BC), were generally 30-minutes in duration. During the interviews, participants were asked about knowledge, perceptions, access, and uptake of the COVID-19 vaccine (see Appendix A for the interview guides for each group). Interviews were undertaken between July 2021 and January 2022. The ON batch began on July 8^th^, 2021, prior to the implementation of mandatory COVID-19 vaccination, and was completed by November 13^th^, 2021. The BC batch began on September 20^th^, 2021, and was completed by January 30^th^, 2022. We included a range of data collection dates to capture the temporal backdrop to the shared experiences of the research participants. It was important to capture these perspectives across and within the milieu of changing mandates and public health protocols. Participants were recruited via community engagement events, vaccination centers, social media, word-of-mouth, and snowball sampling.

The concept of “information power” was used to guide the determination of sample size, where interviews were conducted with consenting participants who had personal or staff experiences with COVID-19 vaccine uptake, access, and confidence until enough information was present to adequately achieve our objective (Malterud et al., 2016). Language supports were offered to all participants, but based on participant preference, all interviews were completed in English.

### Analysis

Interviews were audio recorded and transcribed verbatim. Data were anonymized and all participants were provided with a coded identity. Interview transcripts were analyzed manually through the principles of open, focused, and thematic coding (Braun & Clarke, 2013). Graduate student researchers (BM, DD, RS, ZK) coded each interview independently and in duplicate, meeting regularly with SK to discuss progress, insights, and next steps. The team held two discussions with the broader qualitative working group (GW, DB, SAL, SSA) to iteratively review data analysis, preliminary findings, and confirm information power (Malterud et al., 2016).

### Rigor

Trustworthiness and credibility were achieved through prolonged analytical engagement, the generation of rich descriptive accounts, and peer checking (Lincoln & Guba, 1986). The data analysis team met weekly to discuss biases related to allopathic treatments, reflections on the data, impressions from conducting the interviews and/or listening to the transcripts, and directions for data analysis.

### Positionality

The lead author of this paper (SK) is a South Asian woman raised in the GTA and is currently a postdoctoral researcher. She completed her PhD studies working with South Asian pregnant people and has diverse experience working with different qualitative and mixed-methods methodologies and conducting interviews in many different contexts and with different populations. BM, DD, RS, and ZK each have graduate-level training in qualitative research and currently live and work in Southern Ontario. BM, DD, and ZK also identify as South Asian. Authors RN, DB, and SAL live and work in BC, bringing diverse qualitative research and COVID-19 vaccine access experience to the discussion. GW (pediatrician), RdS (epidemiologist), and SSA (internist) brought clinical experience and methodological expertise to this study.

## Results

25 participants (15 from ON and 10 from BC) were interviewed between July 8^th^, 2021 and January 30^th^, 2022. Participants included South Asian community members (n=10), South Asian advocacy group leaders (n=9), and public health staff who work predominantly with the South Asian community (n=6). The ages of South Asian community members ranged from 19 to 69 years old. All public health staff were medical doctors and advocacy group leaders had the following roles: student advocates, vaccine ambassadors, vaccine outreach workers, community organization leads, and health support workers (Table 1). The advisory working group held regular discussions to plan and execute community engagement activities, review preliminary data, and assess information power. Through these detailed discussions, it was decided that 25 participants across groups and provinces were sufficient for data richness.

We developed six themes across the experiences and perspectives of those interviewed as they pertain to risk and vaccine uptake, access, and confidence: 1) Risk perceptions; 2) Sources of trusted information (traditional and non-traditional); 3) Broader life impacts of COVID-19 and the pandemic on individuals, families, and society; 4) Experiences with COVID-19 mandates and policies (including temporal and generational differences); 5) Awareness and outreach; and 6) Language, culture, and context.

### 1) Perceptions of COVID-19 risk

Participants, specifically those who self-described as lacking vaccine confidence, being disinterested in, or refusing the COVID-19 vaccine, discussed their perceptions of COVID-19 risk from the point of view of their health status, personal risk perceptions, and their perceptions of COVID-19 and vaccine risk. Most participants mentioned they led a healthy lifestyle, were in good health, and were free of chronic diseases or illnesses. In describing this, many participants perceived COVID-19 was a serious disease for those with chronic diseases, while those who were healthy did not need to be as worried or concerned, but should remain cautious and follow public health recommendations. They also perceived that COVID-19 was not as severe in children.

Participants discussed their perceptions of COVID-19 as a whole and how information about COVID-19 was reported. For some participants, there was mistrust in how data were shared, with beliefs that COVID-19 transmission statistics reported in mainstream media were exaggerated and COVID-19 was not as serious as it was portrayed. Participants also compared COVID-19 to other illnesses such as the common cold and mild flu. One community member in ON stated: “*How I see it is in a way it’s not really like a huge deadly concern to my age group* [young adult], *but it is susceptible to catch it, to give it, and stuff like that, if that makes sense*.” (*Community member – ON)*

While many advocacy group leaders perceived the COVID-19 vaccines to be a *“saviour*”, some community members who self-identified as lacking vaccine confidence seemed to disagree. Many disagreed with vaccinating children, expressed concerns related to the speed of vaccine development and approval, and were discouraged by how quick the rollout was in ON and BC. For example, one community member commented:

> *“And the other apprehension I have is that none of the drug makers, Pfizer, Moderna, whatever, have taken any liability, they have not taken any liability on any side effects from introduction of the vaccine, which is a little bit alarming to me. I think it was all done very, very hastily”* (*Community member*—BC).

In contrast, public health staff viewed their patients’ risk perception as embedded within complex and overlapping social issues related to access, equitable rollout, poverty, and hunger. These systemic issues are integrated within the bedrock of health and wellness. The COVID-19 pandemic further amplified existing challenges and barriers. For example, one individual commented that many of their clients say, *“I’m not going to die by the pandemic, but I am going to die by hunger*.*” (Public health staff—ON)*.

### 2) Sources of trusted information

During the interviews, participants reflected at length on the processes of accessing and evaluating COVID-19-related information. Participants reported using varied sources of information, including ethnic-based sources, and how they distinguished between accurate and false information.

One community member in Ontario stated that they relied on sources that offered statistical or quantitative data:

> “*I got a lot of my information from Instagram and a lot of social media platforms and stuff like that, and also news outlets as well. A lot of the times I see a lot more statistic-based stuff, like how many people got COVID, how many people died of COVID this day or that day, and then obviously news outlets will also tell symptoms and more information about the virus*.*” (Community member – ON)*

Community members, advocacy group leaders, and public health staff described the high use of ethnic media stations (TV and radio programs conducted in native South Asian languages), social media (e.g., WhatsApp), and word of mouth from family, friends, and coworkers. Participants highlighted generational differences in the methods of obtaining COVID-19/vaccine-related information, as well as the different sources for this information. Specifically, one advocacy group leader commented that younger individuals typically used non-ethnic media to access information, while older individuals were more likely to access and use ethnic media or their family/friends as a valued source of information.

Family members were viewed to be a highly trusted source of information by members of the South Asian community and commonly influenced people’s vaccination decision. For example, one advocacy group leader said, *“Like if people around them aren’t getting the vaccine they’re going to be seeing that view of why they shouldn’t get it. If they’re surrounded by people and all their family members have gotten the COVID-19 vaccine it further nudges them*.*” (Community Advisory Group Leader—BC)*.

Over the course of the pandemic, advocacy group leaders and public health staff participants reported being heavily involved in fostering evidence-informed dialogue with community members. They worked to address misinformation in the community and provide reliable sources of information, especially at vaccine outreach centers.

> “*In the South Asian community, especially within [city], a lot of the elderly population are on WhatsApp. One of the main ways they get their information, my parents included, is just through quick articles being shared throughout WhatsApp. In the very beginning of the vaccine rollout, it wasn’t uncommon to see when you were trying to explain to a patient why the COVID-19 vaccine is safe to get and important to get they would pull out their phone and show a WhatsApp article from whatever site. And then we would have to take the time to explain, okay, this may not be a credible site, or this may not be information that came from a trusted physician or researcher. And we would have to call out one of the culturally competent physicians to come and explain the more complex medical jargon contained within these articles from WhatsApp that the patient may not be able to fully understand*.” *(Community Advisory Group Leader—ON)*

Connections to one’s birth country was also an important source of information for many. Participants discussed that South Asian community members in ON and BC often referred to the state of their home country when determining the seriousness of COVID-19 and reasons for mistrust or confidence in vaccine.

### 3) Broader life impacts of COVID-19 and the pandemic

During the interviews, participants described how COVID-19 affected them physically, mentally, emotionally, and financially. Physically, many participants described the impacts of improved public health measures (personal protection equipment for workers, mask protocols, hand washing) on their overall health. Others described how they were negatively impacted by contracting COVID-19 or by the side effects of receiving the first dose of the COVID-19 vaccine. For example, one community member recalled the following: *“After my first shot, I started getting this sinking feeling around the middle of my chest. I had a heart attack years ago so I should be worried*.*” (Community member –BC)*

Participants also described the burden that COVID-19 restrictions imposed on their mental health and well-being. Public health guidelines and regulations promoting social distancing and quarantining negatively affected many participants. For many, this led to increased stress due to minimal contact with family and friends.

> *“Being a graduate of 2020 in my high school year, I was basically robbed of my senior year, so I was kind of robbed of saying goodbye to a bunch of school friends that I haven’t seen since March basically. And also, university, especially first year, was very, very difficult for me and very stressful for me. That’s where I’d say COVID-19 mainly impacted it was my first year because it was remote and because no one really helped you specifically. Everyone was kind of doing their own thing. And then that was very stressful, and all the workload was also piling up and everything too*.*” (Community member – ON)*

Others commented on the long-term mental health impacts of COVID-19 restrictions. Advocacy group leaders and public health staff described at length the need to collaborate with diverse organizations to raise funds to support youth mental health campaigns. One participant discussed the negative mental and emotional impacts of the divisive public opinions on the COVID-19 vaccine:

> “*That’s another thing that really upset me about the vaccine is that it really caused a divide. People are passionately for and passionately against and it really causes a divide, which is really sad that I’ve experienced that in my life*.*” (Community member— BC)*.

Others described large-scale financial strains and how that resulted in mental health impacts:

> *“We’ve seen mental health worsen, we’ve seen other health conditions be affected due to delays, we’ve seen incomes, people losing their homes, there’s been so many different businesses closing. All these hardships that people are facing, and of course we know that stress is an independent risk factor for poor health outcomes, anxiety, and stress*.*” (public health staff – ON)*.

However, financially, there were mixed experiences on the impact of COVID-19 among participants. Some were financially stable during the pandemic and did not notice any changes or strains on their financial health. However, others were financially impacted when they were unable to work due to contracting COVID-19 or due to loss of employment. This was further exacerbated by ineligibility for financial aid. On the other hand, a few community members were critical of the Canadian government’s approach to pandemic aid as it related to the supports offered to small business owners.

### 4) COVID-19 Guidelines and Policies

Participants described their attitudes as well as general observations regarding the attitudes of the family and friends towards COVID-19 mandates and policies. This presented itself as varied opinions on public health mandates and restrictions. Several participants stated that they adhered to and agreed with COVID-19 mandates and policies, such as masking and social distancing. However, participants also questioned inconsistent COVID-19 mandates and policies across different settings and media outlets. For example, one participant expressed their frustration with public education efforts in reference to mask mandates. Participants also described how they perceived vaccine mandates and policies to be contradictory and unfair. Several believed people should be allowed a choice without ‘carrots’ being dangled in their view or ‘sticks’ being used by workplaces. A few participants discussed the contradictory nature of vaccine mandates:

> *“Government shouldn’t impose this vaccine to the people who didn’t want it. They are trying to limit the people who go to hospital, I agree with that—and the restaurant and theatre. But they are not [limiting] the people who didn’t get the vaccine, like me to go into any store. The people who are visiting the theatre are also visiting the store as well. What kind of policy this is? It doesn’t look like fair. So, I don’t know what they are doing*.*” (Community member—BC)*.

Another participant described how COVID-19 ‘return to work’ policies differed by workplace setting, causing frustration. Though the vaccine mandates and policies were viewed as contradictory in some instances, the overall message was that they were effective in increasing vaccine uptake within the South Asian community.

> *“However, once the stick came out, like the workplace mandates and you can’t go to a gym without getting vaccinated, stuff like that, the more serious ones like the essential ones. Then I would say it would go from 10% of the patients we saw vaccinated to 60%. Most of the people coming in were due to the mandates*.*” (Advocacy group leader—ON)*

In addition, a few participants discussed generational differences in the South Asian community’s attitudes towards COVID-19 stay-at-home policies and vaccine mandates. Some found members of the older generation were more relaxed about the guidelines. For example, one advocacy group leader said:

> *“…what we find is that the elderly especially because we are a temple, God’s place and you don’t get sick at God’s place, you come here to get well. So, it’s hard for us to tell them to stay home if you’re not feeling well, they want to come and pray so that they feel better. So, that’s a kind of conflict between what should be done, what the health department is saying and what people feel about it, but the younger generation is more accepting the guidelines and following them.” (Advocacy group leader -BC)*

Another advocacy group leader stated the senior population trusted medical interventions more than younger individuals. For example, a participant explained that views on COVID-19 vaccines appeared to be generational, with *“the elderly, coming from the generation they do, have complete 100% trust in the vaccine and the health system whereas the views of younger individuals varied*.*” (Advocacy group leader –BC)*,

### 5) Awareness and Outreach

Public health staff and advocacy group leaders discussed the significance of community engagement, as well as partnerships and collaboration, in the context of vaccine uptake, provision, and education. Participants described the importance of engaging with the community through both traditional (local news stations) and non-traditional media sources (radio, ethnic television stations, print media, South Asian networks (i.e., Connect FM, Omni, Sher-E-Punjab), and social media) to target barriers in vaccine education and provision to meet people where they are. One participant stated engaging with the community demonstrated there was “*a definite knowledge translation issue” (Public health staff—BC)*. Another participant said the following on the importance of engagement:

> *“…we live in a very reactive society. … I think we should equally spend time and energy in developing structures in prevention and health promotion. And case in point, COVID is a role of self-realisation and self-management of chronic diseases…the Public Health Agency of Canada, the Innovation Canada, should look at creating very engaged and institutional trusted multi-cultural, multi-ethnic diversity programmes which are effective.” (Public health staff—BC)*.

Further, participants described how they used partnerships and collaborations to effectively facilitate vaccine education and provision. Two Public health staff were also members of a South Asian collective of physicians aiming to improve vaccine access, education, and uptake. They discussed how they partnered with Facebook and other grassroots organizations, as well as influencers and community leaders (i.e., politicians, celebrities, athletes) to address knowledge gaps and universal concerns. One participant said the campaign included *“people of all different ethnic backgrounds and different professions advocating and speaking in favour of the vaccine so that people had role models (Public health staff—BC)*.” Another participant described having influencers from the community, such as local politicians and South Asian politicians, as a *“power of strength (Public health staff—BC)*.”

Overall, the importance of meeting people where they are was a common theme connecting the experiences of public health staff and advocacy group leaders in vaccine education, provision, and uptake. This included addressing potential barriers at vaccine clinics, such as language, advocating to bring vaccines to the people, as well as facilitating vaccine education and amplifying public health messaging through various media sources the South Asian community engages with and by going to places of worship. For example:

> *“I saw the challenge, our team saw the challenge, was the availability of vaccine at the right time, at the right place, for the right people. And to define that, has been a challenge because [public health] would like to do a vaccination in a designated clinic, and they want people to come there rather than the vaccine goes to where people are*.*” (Public health staff—BC)*
>
> *“I feel like one of the best things we’re doing right now is the mobile outreach. It’s very difficult to get reliable healthcare information out to the most vulnerable populations. So, I feel that instead of relying on them to come to us, us going to them in places where especially elderly populations who are not technologically savvy, who don’t really watch the news or read up on articles” (Advocacy group leader –ON)*

### 6) Language and cultural context

Many public health staff and advocacy group leaders described language and cultural context as being rooted in their work in vaccine provision and education, as well as strategies to improve vaccine uptake. In response to how they tailored their approach to vaccine education and provision in different populations, participants said the following:

> *“…[The healthcare facility] wanted some vaccine clinics in our Gurdwaras, which are our places of worship. Then we had meetings to help support [the Gurdwara], followups, see how we could do it better, make sure it was being done in a culturally safe and acceptable manner*.*” (Public health staff—BC)*
>
> *“Honestly, the questions everyone has are universal, the concerns everyone has are universal, it’s just how you’re addressing them. It’s basically addressing them so that they’re culturally and language appropriate. So, I can go on mainstream media and talk about the same thing and then I can go on OMNI Punjabi and talk about the same thing, there is absolutely no change in my messaging and there is no change in the questions people have. It’s just how I’m communicating. When you see someone who looks like you and speaks like you, you will listen*.*” (Public health staff—BC)*

Several participants emphasized the importance of translation and representation after observing language was a consistent barrier to vaccine provision in terms of registration and education (having questions and concerns addressed). For example, one physician said, *“In a multi-ethnic and multi-cultural society, language is very important*.*” (Public health staff—BC)*

Some participants not only mentioned language as a barrier, but also having a trusted South Asian healthcare worker who was able to relate to community members, emphasizing the importance of cultural matching between physicians and community members:

> *“… so I think a major barrier, especially when we’re talking about immigrant populations, language barrier is a big one. Finding a doctor who can actually have a good conversation and can explain things in medical terms is always very tough, which is one of the benefits that we have here actually because we have a lot of Punjabi-speaking doctors here” (Public health staff—ON)*

Another participant said the following in response to COVID-19 education barriers, reiterating the importance of relaying information in preferred languages instead of leaving the responsibility of translation with media staff or community members:

> *“I feel like, and this is one I actually still see is out there, that we were always chasing information. And what I mean is that when information was coming up from authorities, whether it’s the government or from the health authority, it was always coming out in English and the translation was left to the community to do so. A lot of the education, you were relying on whether the reporters or media anchors to actually interpret what the briefing was saying, whether written or in person, and then understand that and then present it to the community. Inherently, in that multi-step process, things get lost in translation, the meaning can get altered. And so now, what I have seen, some of the briefings online, they will post in Punjabi at the same time and that was not being done*…*but that was something that I think would go a long way*.*” (Public health staff—BC)*

A few participants discussed South Asian representation in terms of addressing questions and debunking myths that were usually addressed in English. One participant said that representation *“was lacking and that misinformation was spreading*” (*Community Advisory Group Leader –BC*). However, they also mentioned several members of the South Asian community stepped forward to help address concerns and support vaccine uptake.

## Discussion

Through a comprehensive analysis of the study data, we identified six themes across the experiences and perspectives of participants from Ontario and British Columbia as they pertain to vaccine uptake, access, and confidence: 1) Risk perceptions; 2) Sources of trusted information; 3) Broader life impacts of COVID-19 and the pandemic on individuals, families, and society; 4) Experiences with COVID-19 mandates and policies (including temporal and generational differences); 5) Awareness and outreach; and 6) Language and cultural context. Vaccination outreach programs that are tailored and rooted in information that is in native languages and builds upon cultural contexts and understandings can be successful in engaging socially marginalized sub-sets of the population. These beliefs require exploration and understanding to develop the many dynamic threads of a successful pandemic response.Many people who experience vaccine concerns may still choose to engage in conversations about COVID-19 vaccine safety, efficacy, and necessity (Mills et al., 2020), which makes it vital for outreach efforts to meet individuals where they are.

Although most South Asians living in Canada have been vaccinated in major Ontario and British Columbia urban centres, some issues related to limited confidence because of long-term impacts/side effects, preference for alternative therapies, and mistrust in institutions, governments, and pharmaceutical companies are still observed (Office for National Statistics, 2021). “19 to Zero”, an interdisciplinary coalition, analyzed general COVID-19 social media trends using deep-learning modeling of natural language processing. The group demonstrates that, in general and consistently since November 2020, low vaccine confidence is fueled by mistrust in institutions, government, pharmaceutical companies, and health authorities, and emphasizes the political nature of COVID-19 vaccination (19 to Zero Research, 2021). Concern for vaccine safety is usually the second most important topic, and vaccine confident networks are mostly concerned about roll-out and accessibility (19 to Zero Research, 2021). Data from the COVID CommUNITY study describes the top three most trusted sources of information related to COVID-19 being health care providers and public health, traditional media sources, and social media (Anand et al., 2022) parallels the descriptions provided by participants included in this qualitative exploration.

An important component of this research study is the kit of tools and recommendations that can be applied to future public health programs, initiatives, and knowledge translation activities. These include rooting outreach activities with attention to language, cultural matching between physicians and community members, and local leadership that is representative of the communities of interest. The most effective interventions for addressing vaccine confidence are iterative and multi-pronged strategies that are community-, culturally-, and language-focused (Jongen et al., 2017; MacDonald et al., 2017; Schoch-Spana et al., 2021). Deep engagement has been shown to improve the promotion and implementation of vaccine programs, resulting in a >25% improvement in vaccine uptake (Hussain et al., 2022; Jarrett et al., 2015). This includes working collaboratively with religious or influential community leaders to foster trust, deliver messages, and facilitate community engagement (Hussain et al., 2022). The engagement of primary care providers with communities through outreach with tailored messaging has also been shown to be effective in improving vaccine uptake (Jarrett et al., 2015; Bogart et al., 2021; Lieu et al., 2022). In addition, addressing contextual influences and tailoring messages to specific beliefs and concerns to inform outreach rooted in community-engaged approaches (e.g., co-designed materials that are written first and foremost in native languages) was demonstrated to be integral in vaccination programs (Schoch-Spana et al., 2021; Jarrett et al., 2015; AuYoung et al., 2022; Marcelin et al., 2021).

The aforementioned points can provide scaffolding and recommendations for the implementation of successful programs now and in future pandemics:

1. Public health programs must take a strengths-based approach that builds upon historical sources of mistrust/experiences and cultural-linguistic needs
2. Community partnerships are key to guiding and making recommendations on how to best engage community members (including avenues for communication such as social media platforms and traditional media)
3. Key pieces of information included in vaccine outreach must address individual risk perceptions and acknowledge broader impacts of pandemic decision-making (e.g., mandates)

The strengths of this research include: 1) the involvement of diverse stakeholders and community members; 2) using community-engaged research practices, allowing us to tailor the study approaches to best meet the needs of the target population and to create safe and culturally appropriate research activities; 3) applying qualitative methods, as part of a larger series of studies, to capture a detailed review of experiences and perspectives of COVID-19 vaccination in a population that has historically faced barriers to engagement in healthcare and research.

The limitations of this research include: 1) the potential that attracting a larger or more diverse range of stakeholders may have yielded additional insights; 2) a focus only on South Asian communities residing in large urban centres—community members living in rural and remote areas may have held different perspectives or had differing experiences with COVID-19 vaccine uptake, confidence, and access. Future research could aim to capture the aforementioned two areas to further deeper and strengthen the conversation within this space.

## Conclusion

We identified six themes across the experiences and perspectives of participants from Ontario and British Columbia as they pertain to vaccine uptake, access, and confidence: 1) Risk perceptions; 2) Sources of trusted information; 3) Broader life impacts of COVID-19 and the pandemic on individuals, families, and society; 4) Experiences with COVID-19 mandates and policies (including temporal and generational differences); 5) Awareness and outreach; and 6) Language and cultural context. Awareness, outreach, and being sensitive to the application of language, culture, and context may help improve the success of vaccination programs. An opportunity exists to foster culturally authentic vaccine programs that address the direct concerns of key stakeholder groups and work to address ongoing and worsening health inequities.

## Supporting information

Appendix A (Interview Questions)

## Data Availability

All data produced in the present work are contained in the manuscript.

## Contributions to Knowledge

- Using qualitative methods, we sought to explore the perceptions of COVID-19 risk, vaccine access, uptake, and confidence among South Asian Canadians.
- We identified six themes as they relate to vaccine uptake and confidence: 1) Risk perceptions; 2) Sources of trusted information (ethnic and non-ethnic sources); 3) Broader life impacts of COVID-19 and the pandemic on individuals, families, and society; 4) Experiences with COVID-19 mandates and policies (including temporal and generational differences); 5) Awareness and outreach; and 6) Language and cultural context.
- Understanding factors and strategies that build vaccine confidence can guide our approach to increase vaccine acceptance in the current COVID-19 pandemic as well as future pandemics.

