## Supplementary material for "Perceptions of COVID-19 risk, vaccine access, and confidence: a qualitative analysis of South Asians in Canada": Table 1

| Characteristic | Value |
| --- | --- |
| Total # living and working in:  ONTARIO  BRITISH COLUMBIA | 15  10 |
| Total # of Community members  Median age  # Males  # Females | 10  32 (Range: 19-69)  5  5 |
| Total # of Advocacy group leaders  Vaccine advocates or ambassadors  Executives or leaders in local organizations  Other (e.g., general health program coordinator) | 9  4  3  2 |
| Total # Public Health Staff  MD certification | 6  6 |

**Table 1. Demographic characteristics of participants enrolled**
