## Appendix A (Interview Questions) for "Perceptions of COVID-19 risk, vaccine access, and confidence: a qualitative analysis of South Asians in Canada"

**1. Participant Interview Questions**

Thank you for participating in the COVID CommUNITY Study. We would like to learn your about your knowledge, perception and attitudes towards COVID-19 vaccination. We have a set of questions to ask you. We will record this interview, however, your comments will be kept confidential and no personal information such as name, age or birthday will be recorded.

1. Tell me briefly a little bit about yourself and your current health.
   a. Age, gender, current health, household structure?
2. What do you know about the COVID-19? Who gets it? What are the health consequences? What measures can you take to protect yourself?
3. How has the pandemic affected your life (e.g. physical/mental health, financial health, lifestyle, religious practices, family interactions)?
4. What do you know about the COVID-19 vaccines?
   a. Prompt: What are your thoughts on the different COVID vaccines (e.g. Pfizer, Moderna, Astra Zeneca)?
   b. Prompt: What are your views on COVID-19 vaccine side-effects?
   c. Prompt: What are your thoughts on the COVID vaccine for children?
5. When you think of the COVID-19 vaccine, what comes to mind? What do you find the most concerning? The most reassuring?

a. Prompt: think back to the time before you received your first dose?

6. What views do people in your community, family & friends have regarding the COVID-19 vaccines?
7. Tell me about a time that you made a decision that involved weighing the risks of the COVID-19 vaccine. What did you decide? How did you come to that decision?
8. Are there any barriers to getting your COVID-19 vaccine? (e.g. transportation, time of day, time off work, etc)
9. How do you get information about the COVID-19 and the vaccine? What sources do you trust and what ones do you distrust? (e.g. Government websites, family/friends, my family doctor, community/spiritual leaders, healthcare professionals, family, etc.)
10. How do you distinguish between real and false information?
11. Is there anything else that you feel we should know about your experience?

12. Vaccine mandates

**2. Public health staff – Interview questions**

1. Tell me about yourself and your role in public health?
   a. How have you been involved in the provision of healthcare services related to COVID-19?
2. Please tell me about your experiences of vaccine education and provision
   a. How have you delivered education about vaccines?
   b. What barriers have you experienced or witnessed?
   c. What has helped in the education and delivery of vaccines?
3. When reflecting upon your experiences so far, how has vaccine education and provision been targeted across diverse populations?
   a. How have you tailored your approach to different populations?
   b. How have you experienced practicing with populations that face barriers to engagement?
      i. Prompt: Language barriers, access barriers, cultural barriers, etc
         1. What have been successful solutions to barriers?
         2. Pandemic burnout, paid sick days,
4. Please tell me about your experiences of supporting and working with members of South Asian communities
   a. Are there any barriers that may be experienced related to COVID-19 education?
   b. Are there any barriers that me be experienced related to vaccine education and uptake?
   c. Are there any enablers for COVID-19 education?
   d. Are there any enablers for vaccine education or uptake? e.g. community partnerships, existing or new
5. Have you adapted your practice to reflect the needs of members of South Asian communities?
   a. What gaps have you observed?
6. Tell me about your observations around vaccine-related knowledge and update in members of the South Asian community.
   a. Where do you think that community members access their information?
   b. What can healthcare providers do to address issues of COVID-19 vaccine hesitancy?
   c. How might you think the opinions of others (e.g., family members and community leaders) influence decision-making or perspectives related to COVID-19 vaccination?
7. Is there anything that we haven’t asked about that you think is important for us to know?

**3. Community advocacy group leaders – Interview questions**

1. Tell me about yourself and your organization?
   a. How have you been involved in the provision of services related to COVID-19 education or vaccination?
2. Please tell me about your experiences of supporting and working with members of your community and organization?
   a. Are there any barriers that may be experienced related to COVID-19 education?
   b. Are there any barriers that me be experienced related to vaccine education and uptake?
   c. Are there any enablers for COVID-19 education?
   d. Are there any enablers for vaccine education or uptake? e.g. partnerships with government organizations? Existing or new partnerships?
3. When you think of the COVID-19 vaccine, what comes to mind? What do you find the most concerning? The most reassuring?
4. What views do people in your community have regarding the COVID-19 vaccines?
5. Tell me about your observations around vaccine-related knowledge and update in members of the South Asian community.
   a. Where do you think that community members access their information?
   b. What can healthcare providers do to address issues of COVID-19 vaccine hesitancy?
   c. How might you think the opinions of others (e.g., family members and community leaders) influence decision-making or perspectives related to COVID-19 vaccination?
6. Please tell me about the experiences of your community around the COVID vaccines?
   a. How has your organization been involved in education about vaccines?
   b. What barriers have you experienced or witnessed?
   c. What has helped in the education and delivery of vaccines?
7. Is there anything that we haven’t asked about that you think is important for us to know?
